# Protocol for a randomized cross-over study measuring the effect of reduced protein intake on autophagic flux in healthy adults

**DOI:** 10.1101/2024.06.16.24308986

**Authors:** Célia Fourrier, Leonie K. Heilbronn, Xiao Tong Teong, Jemima R. Gore, Timothy J. Sargeant, Julien Bensalem

## Abstract

Autophagy is a cellular mechanism that degrades damaged or unwanted material from cells and is particularly important during ageing. Autophagy has been widely studied in pre-clinical models and is known to respond to nutrient availability and in particular amino acids. However, clinical data are limited. This protocol paper describes a randomized cross-over clinical study investigating the effect of a four-week long reduction of dietary protein intake on autophagic flux (autophagic degradative activity) measured via a blood test in healthy adults. Sixty-one healthy participants will be recruited. Study participants will be randomly assigned to one of two diets for four weeks, then cross over to the other diet with a four-week washout period between diets. The diets will be: 1) Average Protein Diet: a control comparator diet with a macronutrient profile of 20% of energy from protein, 35% from fat (<10% saturated) and 45% from carbohydrate, with >30 g of fiber/day; 2) Reduced Protein Diet: 10% protein, 35% fat (<10% saturated), 55% carbohydrate, >30 g of fiber/day. This diet will provide 0.7 g of protein per kg of body weight (the recommended minimum daily protein intake). The primary outcome will be the change in autophagic flux between each diet. This study aims to determine whether reducing dietary protein intake in healthy adults can increase autophagy in humans. If positive, it creates the possibility that nutritional strategies could be used to prevent or delay autophagy-related diseases such as Alzheimer disease or atherosclerosis.

## Background

Autophagy is an intracellular process that collects and degrades unwanted or damaged organelles and macromolecules and recycles the degraded material to replenish macronutrients during nutritional stress. Autophagy is critical for healthy cellular function and the prevention of age-related disease (1). Autophagy therefore represents an important target for treatments that seek to prevent or delay age-related disease. Amongst potential mediators of autophagy, nutrient restriction has been shown to induce autophagy in *in vitro* and *in vivo* studies (2, 3), notably through inhibition of the mechanistic target of rapamycin complex 1 (mTORC1) (4). Reduced protein intake was shown to reduce mTORC1 activity (5, 6) and increased carbohydrate:protein ratios were associated with increased lifespan in animals (7), and is correlated with reduced risk of death in people aged 50-65 and reduced risk of cancer and diabetes mortality over an 18-year follow up (8). Protein consumption in humans increased mTORC1 activity in monocytes and changed LC3-positive puncta abundance, consistent with a decrease in autophagy, although autophagic *flux* itself was not assessed in that study (9). Consistent with these data, it has been shown that high protein diets increase mTORC1 activity and decrease lysosomal system function to exacerbate a model of heart disease (9, 10).

It is important to note that measuring autophagic degradation activity, named autophagic flux, is particularly challenging in humans. Indeed, existing methods require either genetic manipulation (e.g. a transgene coding for a fluorescent probe) (2, 11, 12) or the use of a lysosomal inhibitor such as chloroquine or bafilomycin to block degradation of autophagic material by the lysosome (12, 13). Amounts of LC3-II, a marker of autophagic vesicles, can be measured to determine how much autophagic material should have been degraded during the lysosomal inhibition period in comparison to a non-treated paired sample. To study autophagic flux in humans, we have developed a method where the lysosomal inhibitor chloroquine is added to whole blood from fasting participants, and autophagic flux is measured in peripheral blood mononuclear cells (PBMCs) isolated from these treated blood samples (14). LC3-II quantification is performed by enzyme-linked immunosorbent assay (ELISA). This method measures autophagic flux from cells in a physiological environment (participant’s blood) in comparison to cells that would be cultured in a nutrient-rich artificial culture medium which could itself alter autophagy (2, 15, 16).

The aim of this randomized cross over trial is to establish whether a four-week-long reduction of dietary protein intake increases autophagic flux in healthy adults as compared to a moderate protein intake, approximating habitual diets in Australia (17).

## Methods and design

### Ethics and study registration

The Human Research Ethics Committee (HREC) of the University of Adelaide on 31^st^ August 2021 (HREC reference number: H-2021-154, current version at submission of this paper: v5.2 dated 30^th^ May 2024) gave ethical approval for this work. Serious adverse events considered to be related to the study will be reported to the HREC by the PI.

The study was registered on anzctr.org.au with the following identifier: ACTRN12623000260628 and entitled ‘*Effect of Nutritional INterventions on Autophagy: The NINA study*’. The NINA study is a single-site study performed at the South Australian Health and Medical Research Institute (SAHMRI) in Adelaide, South Australia, Australia, by researchers from SAHMRI and the University of Adelaide.

### Study design

The NINA study is a single-site, single-blinded, randomized cross-over study.

### Intervention masking

Given that the NINA study is a diet intervention study, the investigator interviewing the participants and providing the diets will not be blinded to the group assignments. However, the laboratory work, data management and final analyses of the primary outcomes will be carried out by technical investigators who are blinded to each participant’s allocated group. Participants will be blinded to dietary interventions. To prevent study participants from trying to guess which diet is the ‘Average Protein Diet’ and which diet is the ‘Reduced Protein Diet’ and therefore avoid bias, the participant information sheet does not outline the purpose of the study which is to investigate the effect of dietary protein intake on autophagy (**Additional file 1**). The wording has been kept general and states that the study aims to measure the effect of diet composition on autophagy in humans. The macronutrient profile of the diets will be manipulated with the assistance of a high or low protein supplemental powder to keep the diets as similar as possible and to mask the identity of the diets as much as possible.

### Participants

Participants are recruited from the Adelaide metropolitan area (South Australia, Australia). The study is advertised through SAHMRI and The University of Adelaide networks and on social media.

Participant inclusion and exclusion criteria for the study are listed in **Table 1**.

**Table 1.**
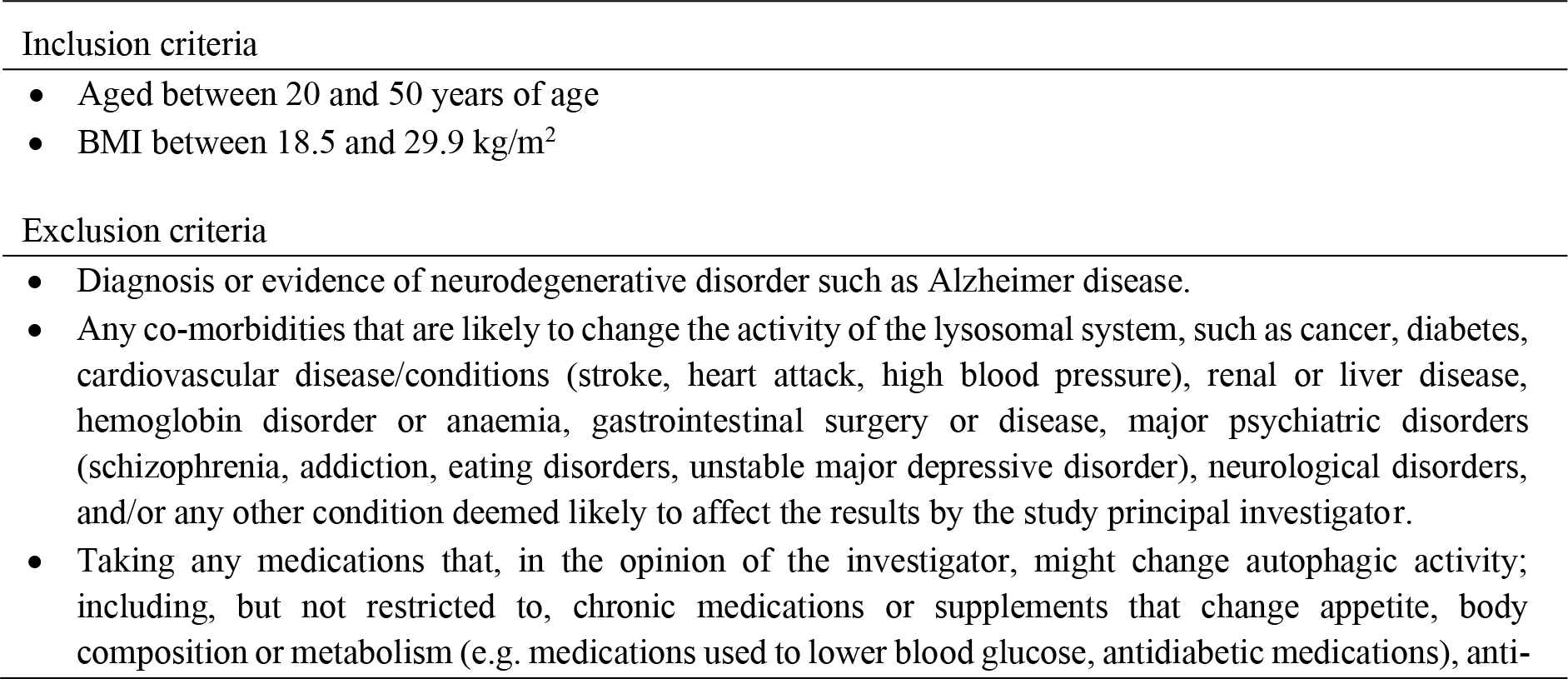
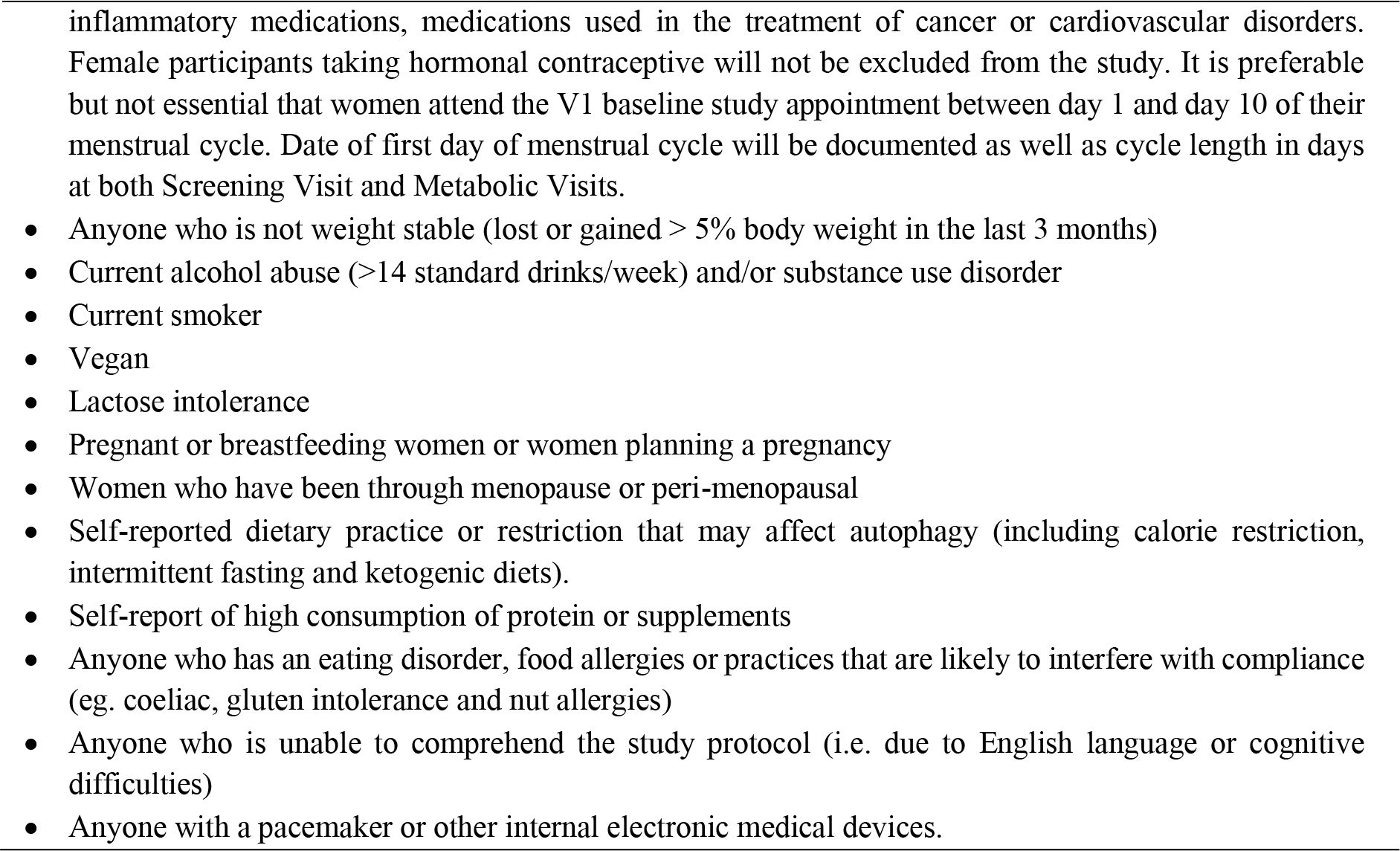
Participant inclusion and exclusion criteria.

### Screening

Interested participants are invited to complete an online screening questionnaire using REDCap (Research Electronic Data Capture) which gathers demographic and health-related information, including diet, medical, and exercise history, to assess their eligibility for the study (**Table 1**). At this time, volunteers will also be asked to complete the Night Eating Diagnostic Questionnaire (NEDQ) (18) via REDCap to assess the presence of night eating syndrome which would exclude them from the study. Participants are also provided with a copy of the participant information sheet (**Additional file 1**) and allowed as much time as they require to decide whether they would like to participate.

Potentially eligible participants will be invited to attend SAHMRI and have the research protocol explained to them in detail. Informed consent to participate in the study, including a verbal indication that they understand the general study protocol and requirements will then be obtained (**Additional file 1**). During this session, weight and height will be obtained using standardized procedures to calculate body mass index and ensure suitability for inclusion into the trial. Participants will be asked for the duration of the study not to change their exercise. Baseline exercise level will be measured at screening using the International Physical Activity Validated Questionnaire (IPAQ). Participants will also be asked to complete a 3-day habitual food diary prior to starting the study diet to get an indication of their usual eating habits for use in the analysis. If participants meet the eligibility criteria, they are invited to continue in the remainder of the study. Participants are free to withdraw from the project at any time.

### Randomization

Randomization of participants will be stratified by sex into one of the two study diets at baseline with a ratio of 1:1. The allocation sequence will be determined by a computer-generated random number sequence, using a permuted block design. The randomization schedule will be built into REDCap and unable to be manipulated by research staff.

### Study intervention

Study diets are designed by a research dietitian using FoodWorks Professional (version 10; Xyris Software (Australia) Pty Ltd., Australia). To aid compliance, participants will be provided with individualized 7-day rotating menus to accommodate personal preferences (e.g. cultural preferences, food intolerance, vegetarian) with most of the food provided. Participants will also be asked to complete food checklists daily, recording any food prescribed that was not eaten, and any food eaten that was not prescribed, to be analyzed for compliance. Participants will not be informed which diet is the intervention diet or the control diet.

Study participants will be randomly assigned to one of the two diets for four weeks, before crossing over to the other diet, with a four-week washout period between diets.

*Average Protein Diet*: a control comparator diet that is similar to the ‘typical Australian diet’ will be delivered at calculated energy balance based on equations we have previously validated (19). This diet consists of 20% protein, 35% fat (<10% saturated), and 45% carbohydrate, with >30 g of fiber daily. This group will receive the same information and study visits as the other group.

*Reduced Protein Diet*: this is the intervention diet and will be similar to the *Average Protein Diet*, except that macronutrients will be given at 10% protein, 35% fat (<10% saturated), and 55% carbohydrate, with >30 g of fiber daily. This diet will provide 0.7 g of protein per kg of body weight (the recommended minimum daily protein intake).

The macronutrient profile of the diets will be manipulated with the assistance of a high or low-protein supplemental powder to keep the diets as similar as possible to enable masking of the diet groups. The supplemental powders are commercially available products. To retain masking, they have been repackaged into silver foil pouches that are devoid of information. The powders are dairy-based; because side effects linked to lactose intolerance could arise, lactose intolerance is included as an exclusion criterion. Menus will be designed to allow options for the method of consumption of the powder, but generally the powder will be consumed mixed with a liquid (e.g. milk or water) as a drink. Most of the food will be provided but study participants will be asked to buy some perishable grocery items (eg. fresh milk, juice, fruits, and vegetables).

To prevent study participants from trying to guess which diet is the ‘Average Protein Diet’ and which diet is the ‘Reduced Protein Diet’ and therefore avoid bias, the participant information sheet does not outline that the purpose of the study is to investigate the effect of dietary protein intake on autophagy. The wording has been kept general and states that the study aims to measure the effect of diet composition on autophagy in humans.

### Study visits

Participants will attend four visits in addition to the screening visits. Two visits will take place at the beginning of each diet (V1 and V3), and two visits at the end of each diet (V2 and V4). The study flow chart is illustrated in **Figure 1**.

**Figure 1.**
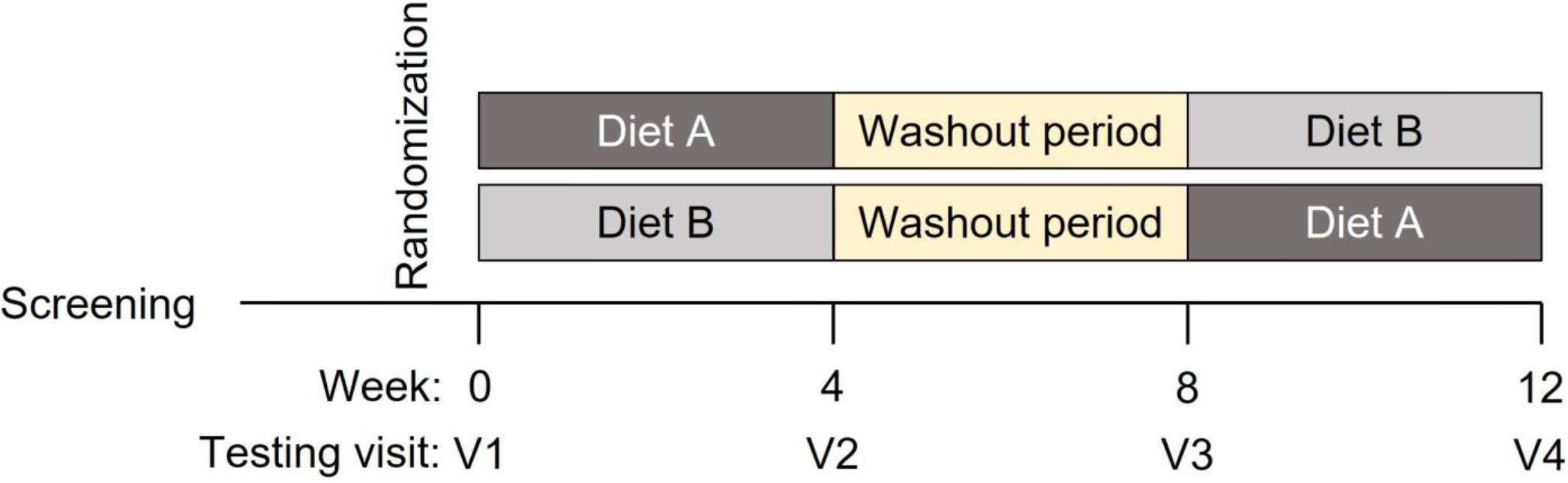
Experimental design of the NINA study. Diet A or Diet B represent the Average Protein Diet and the Reduced Protein Diet, respectively. The randomization will determine the order A-B or B-A.

Participants are asked to fast (including from alcohol and caffeine) and refrain from strenuous exercise for 12 hours overnight prior to a study visit. Participants attend the study visits at SAHMRI North Terrace Adelaide. Study visits are booked in the morning, usually between 7:30 – 9:00 am, at the participants’ convenience.

Eligible participants will attend a first visit (V1) where they will be randomly assigned to one of the two study diets. They will be given diet instructions before starting the intervention. During the first four weeks of the intervention phase, they will receive a 7-day repeating menu plan and matching menu checklists. There will be a short mid diet check-in survey automatically sent via REDCap to the participants at day 14 of each diet period to monitor adherence to the diet and flag any issues participants may be experiencing. After four weeks following the first study diet, study participants will attend a second visit (V2). A four-week washout period begins following V2 and participants will be advised to consume their regular diet. At the end of the washout period, participants will attend a third visit (V3) and be allocated to the diet that they were not allocated to during the first four weeks. They will be given the same instructions as during V1. After four weeks following the second diet, they will attend SAHMRI for a last visit (V4).

An additional SPIRIT (Standard Protocol Items: Recommendations for Interventional Trials) checklist and an additional figure have been provided for further details regarding the study of enrolment, interventions, and assessments for the current study (**Additional file 2** and **Figure 2**).

**Figure 2.**
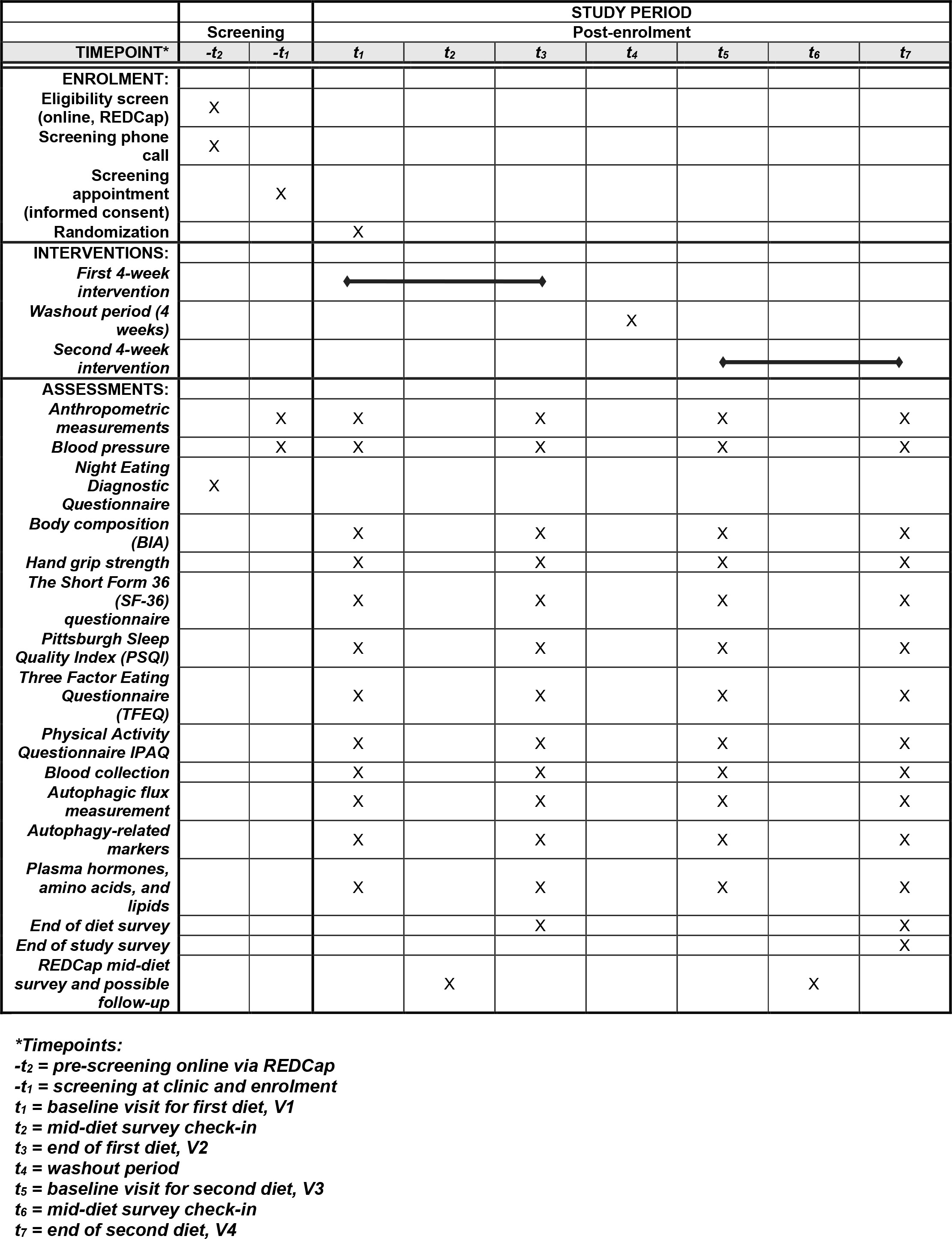
SPIRIT figure. Schedule of enrolment, interventions, and assessments.

#### Data and biochemical samples collected <u>Anthropometry and blood pressure</u>

Height will be measured without shoes to the nearest 0.1 cm using a wall mounted stadiometer. Weight will be measured after voiding, with clothing to the nearest 0.1 kg using a calibrated scale (InBody230, InBody Co). Waist circumference will be measured at the mid-axillary line (halfway point between the lowest rib and the top of the iliac crest), and hip circumference will be measured at the widest circumference of the buttocks using a metal measuring tape. Body mass index (BMI) will be calculated as weight in kilograms divided by height in meters squared. Body composition will be assessed by Bioelectrical Impedance Analysis (BIA) (InBody230, InBody Co). Blood pressure will be measured using an automatic digital monitor and appropriately sized cuff after the participant has been seated for 10 minutes at rest.

#### Hand grip strength

Hand grip strength will be assessed by a digital hand dynamometer (Jamar Plus, Serial number 96179, Performance Health, S.I. Instruments Pty Ltd).

#### Blood collection

Fasting blood samples (up to 60 mL) will be collected by venipuncture and processed immediately or within 30 minutes.

#### Autophagic flux measurement

Autophagic flux will be measured according to the method developed by Bensalem et al (14). Briefly, 6 mL of each blood sample is split into two tubes: one tube will be treated with chloroquine (CQ) to block lysosomal degradation of autophagic cargo LC3-II and the other will be left untreated (unblocked); each sample will then be incubated for 1 h at 37°C. PBMCs are then isolated from both tubes and stored frozen at -80°C for subsequent analysis of protein LC3-II cargo accumulation by ELISA using a commercially available kit. Autophagic flux is determined by the difference in the accumulation of LC3-II over time between blocked and unblocked samples (i.e., what should have been degraded during the one-hour incubation) using the equation ΔLC3-II = (LC3-II+CQ – LC3-II-CQ)/hour.

#### Autophagy-related markers

Change in autophagy-related proteins and mRNA will be assessed by western blot and/or RT-qPCR (e.g., phospho-mTOR, phospho-S6 ribosomal protein, phospho-5’ adenosine monophosphate-activated protein kinase [AMPK], sequestosome-1 [P62/SQSTM1], lysosomal-associated membrane protein 1 [LAMP1]). Molecular cell damage (e.g., lipid oxidation, oxidative stress) will be analyzed using commercially available assays. Analysis of nucleic acids will not reveal sequence-level data and will be used to measure transcript abundance (gene expression) only. Genetic data will not be generated or revealed during this study.

Biological material (e.g. plasma, leukocytes) isolated and stored during this project may be used to identify autophagy biomarkers via, but not limited to, proteomic, metabolomic and/or lipidomic analyses. Material may also be used to develop techniques that allow the measurement of autophagy-related markers in a way that permits identification of specific cell types (such as with flow cytometry) or test reagents or proteins (modified or unmodified) that may be added to blood or cells for the measurement of autophagy and lysosomal processes.

#### Plasma hormones, amino acids, and lipids

Plasma will be snap frozen at -80°C for later assessment of blood glucose, insulin, blood lipids, free fatty acids, glucose regulating hormones (e.g., insulin, C-peptide, adiponectin), inflammatory cytokines (e.g., C-reactive protein), and appetite-regulating hormones (e.g., glucagon-like peptide-1 [GLP-1], glucose-dependent insulinotropic polypeptide [GIP], ghrelin) using commercially available kits. Plasma amino acids will be analyzed by mass spectrometry. Lipids (e.g., total cholesterol, high-density lipoprotein, low-density lipoprotein, triglycerides) will also be analyzed.

#### Questionnaires

To investigate whether dietary protein intake impacts mental status, sleep quality, eating habits, quality of life, and changes in physical activity, a series of questionnaires will be administered before and during the study period: the Short Form 36 (SF-36) (20) to assess the quality of life; the Pittsburgh Sleep Quality Index (PSQI) (21) to assess self-rated sleep habits during the last month; the Three Factor Eating Questionnaire (TFEQ) (22) to assess three cognitive and behavioral domains (or ’factors’) of eating - cognitive restraint, disinhibition, and hunger; and the Physical Activity Questionnaire IPAQ (23).

In addition to the questionnaires, a survey will be conducted at the completion of each study diet (V2 and V4) and at the end of the overall study. This will collect qualitative data about study expectations, perceptions, and experiences.

### Sample size calculation

With existing data available to estimate the expected change in ΔLC3-II (i.e. autophagic flux) between diets, we have assumed a change within-subject of 15%. Based on previously observed ΔLC3-II mean of 277.862 ng LC3B-II/mg protein/h and an SD of 90.675 in healthy subjects, assuming a change of 15% within-subject between both diets, α = 0.05 and a power of 80%, requires a sample size of 53 participants based on a paired Student’s t-test (two-tailed). Assuming a 15% dropout rate, we will recruit n = 61 participants.

### Planned outcomes

#### Primary outcome measure

- ΔAutophagic flux

#### Secondary outcome measures

- ΔAutophagy-related markers
- ΔBody weight
- ΔBody composition
- ΔBlood metabolic markers [lipids, glucose, insulin, amino acids]
- ΔBlood pressure
- ΔMuscular strength
- ΔQuality of life
- ΔSelf-rated sleep habits
- ΔEating behaviours
- ΔPhysical activity
- Diet satisfaction and preference

### Statistical plan

Outcomes will be analyzed using linear mixed effects models to estimate the effect of diet, expressed as a difference in means with 95% confidence interval and 2-sided p-value. Models will account for potential period effects and baseline measurement. Demographic characteristics will be summarized using descriptive statistics.

### Data management

Upon screening, participants are assigned a unique study-specific identifier that is used on all data collection documentation after confirming their enrolment in the study. All electronic data collection and documentation is stored on a password protected REDCap database on SAHMRI’s secure server and can only be accessed by designated research team members. Any paper copies of data will be stored in a locked cabinet within a security swipe card accessed area in SAHMRI.

Biospecimens are stored at SAHMRI in secured freezers, all of which are monitored by a central alert system 24 hours per day seven days a week.

No Data Monitoring Committee (DMC) will be required as this study does not involve significant safety concerns, risks, or complexity.

The results of the study will be made publicly available via official publication and media release at the study’s conclusion.

## Discussion

This study will indicate whether reducing protein intake could be developed as a nutritional strategy to change autophagy to promote health and prevent autophagy-related diseases such as Alzheimer disease or atherosclerosis.

## Trial status

This study was registered on the 10^th^ of March 2023 at anzctr.gov.au under identifier ACTRN12623000260628. The first participant was randomized into the study on the 11^th^ of May, 2023. At the time of submission, randomization of participants into the study was still open. If any important changes to the study protocol become necessary, where appropriate, the PI will notify the HREC, trial register, and the present journal. At time of submission, the current protocol number was version 5.2 dated 30^th^ May 2024. The study is subject to audit at any time by the HREC.

## List of abbreviations

AMPK: 5’ adenosine monophosphate-activated protein kinase
ANZCTR: Australian New Zealand Clinical Trials Registry
BMI: Body mass index
CQ: Chloroquine
CRP: C-reactive protein
ELISA: Enzyme-linked immunosorbent assay
GIP: Glucose-dependent insulinotropic polypeptide
GLP-1: Glucagon-like peptide-1
HREC: Human Research Ethics Committee
LAMP: Lysosomal-associated membrane protein 1
LC3-II: Microtubule-associated proteins 1A/1B light chain 3B II
mTORC1: Mechanistic target of rapamycin complex 1
P62/SQSTM1: Sequestosome-1
PBMCs: Peripheral blood mononuclear cells
REDCap: Research Electronic Data Capture
SAHMRI: South Australian Health and Medical Research Institute
SD: Standard deviation
SPIRIT: Standard Protocol Items: Recommendations for Interventional Trials

## Declarations

### Ethics approval and consent to participate

The study was approved by the University of Adelaide HREC on 30^th^ of August 2021 (HREC reference number: H-2021-154). Study participants have all study details explained to them in writing and in person before giving informed consent.

The present study is conducted in full cooperation with the World Health Organization’s International Standards for Clinical Trial Registries.

### Consent for publication

Not applicable.

### Availability of data and materials

Not applicable.

### Competing interests

TJS and JB are listed as inventors on a related patent—Australia (Provisional) 2019903187;

2019904822; PCT/AU/2020/050908; United Kingdom GB2603664B; USA 17/637,494.

### Authors’ contributions

CF assisted with the study conception, wrote the first draft of the ethics protocol, and wrote the first draft of the manuscript. LKH conceived the study, assisted with writing the ethics protocol, and obtained funding for the study. XTT assisted with study conception and designed the dietary interventions. JRG assisted with writing the ethics protocol and registered the study. TS conceived the study, assisted with writing the ethics protocol, registered the study, and obtained funding for the study. JB conceived the study, wrote the first draft of the ethics protocol, obtained funding for the study, and wrote the first draft of the manuscript. All authors critically revised the manuscript and approved the final manuscript.

## Data Availability

No data are reported in this manuscript.

## Acknowledgment

Acknowledgment is made to the donors of the Standard Award Program in Alzheimer’s Disease Research grant (A2021040S), a program of the BrightFocus Foundation, for support of this research. We acknowledge the University of Adelaide HREC for reviewing our ethics application.

---

**Additional file 1. Study information sheet.** Detailed study information is provided to participants during screening, before the first study visit, and before written consent is obtained. The study information sheet is followed by the participant consent form.

**Additional file 2. SPIRIT checklist.** SPIRIT checklist listing all the items addressed in the current study protocol.

