## Additional file 1. for "Protocol for a randomized cross-over study measuring the effect of reduced protein intake on autophagic flux in healthy adults"

### PARTICIPANT INFORMATION SHEET & CONSENT FORM (PICF)

#### **PROJECT TITLE:** Effect of **N**utritional **IN**terventions on **A**utophagy (The NINA study)

**HUMAN RESEARCH ETHICS COMMITTEE APPROVAL NUMBER: H-2021-154**
**PRINCIPAL INVESTIGATOR:** Dr Timothy Sargeant

Dear Participant,

You are invited to participate in the research project described below.

**What is the project about?**
This research project is about whether what we eat affects autophagic activity. Autophagy is a cellular process that cleanses your cells and is particularly important as we age. Diet composition, fasting or restricting calorie intake have been at the core of strategies that seek to boost autophagy to prevent cell damage and slow cellular ageing. Despite numerous media flooding the internet advocating the advantages of diets that boost autophagy, our knowledge mostly comes from work done in the laboratory and in rodents, not humans.

To properly answer questions about whether nutritional strategies can change autophagy in humans, our research group has developed a first-ever test to measure autophagy in human blood.

This project aims to measure the effect of diet composition on autophagy in blood, using two different diets designed by a qualified dietitian. This will provide information about whether autophagy responds to nutrition in humans, and if it does, how nutrition could be used to treat or prevent diseases known to be impacted by changes in autophagy such as Alzheimer's disease, cardiovascular disease or cancer.

**Who is undertaking the project?**
This project is being conducted by Dr Timothy Sargeant, Dr Julien Bensalem, Dr Célia Fourrier, Ms Kathryn Hattersley, Ms Leanne Hein, Jemima Gore, Helen Checklin, Gemma Barker and Rochelle Botten. (South Australian Health and Medical Research Institute, SAHMRI); and Prof Leonie Heilbronn, Ms Xiao Tong Tong (University of Adelaide). Additional staff may join the research team to assist with this research.

**Why am I being invited to participate?**
You are being invited to participate as you are aged between 20 and 50 years, with a stable weight and a body mass index (BMI) between 18.5 and 29.9 $kg/m^2$.

**What am I being invited to do?**
**Screening Questionnaire**
An online questionnaire containing questions regarding your diet, exercise, medical and surgical history, and eating habits will determine your initial eligibility.

##### Screening visit

After completing the screening questionnaires, you will be invited to attend SAHMRI for a screening visit to determine whether you are eligible for the study. This visit will take approximately 1 hour. In this time, we will explain the study to you in detail and ensure you understand what is involved if you decide to take part. Once you have completed the consent form, we will perform some routine clinical checks (e.g. measure your height, weight and blood pressure). Results from these tests will determine if you meet the eligibility criteria to be invited to take part in the rest of the study. We will also ask you to complete a questionnaire that records the type and amount of physical activity you do. This questionnaire will be repeated at each study visit and you will be asked not to change your exercise pattern for the duration of the study.

##### Food Diary

In order to record your usual eating habits, you will be asked to download an application to your smart phone to allow you to record a 3-day food diary (two week days and one weekend day) prior to the first Metabolic visit.

##### Randomisation:

You will be asked to follow two different 4-week diets and attend four metabolic visits over the course of the study (See Figure 1 below). At your first metabolic visit, you will be randomly allocated to one of the two study diets. You will follow this diet four weeks, followed by a 4-week ‘washout period’ where you will return to your normal eating habits. Then you will follow the other diet for 4-weeks. The order in which you will follow each diet is random, and you cannot choose which diet you complete first.

##### Metabolic Visits

You will be asked attend four metabolic visits, each metabolic visit will take approximately 1.5 hours and will occur at the beginning and end of each 4-week diet. Your schedule for your metabolic visits will be arranged at your screening visit.

For 12 hours before each metabolic visit, you cannot consume any alcohol or caffeine and we will ask that you refrain from strenuous exercise (eg. high intensity workout, running, cycling etc). You will be asked to come into SAHMRI North Terrace in the morning after a 12-hour overnight fast (water intake will be permitted).

At these visits we will measure your weight, blood pressure, waist and hip circumference, and hand grip strength, which is an indicator of overall well-being. You will be asked to complete some questionnaires that will ask you about your general health and wellbeing, mood, sleep, food intake, hunger and physical activity. We will then collect a blood sample from your arm (up to 60 mL, about 3 tablespoons), which will be used to measure your autophagy activity. We will also measure your body composition using Bioelectrical Impedance Analysis (BIA), this is a short, non-invasive, painless procedure that gives us a measurement of your muscle mass and body fat.

At the end of each study diet, we will ask you to complete a survey about your experience on the diet. At the end of the entire study, we will ask you a few extra questions about your diet preference.

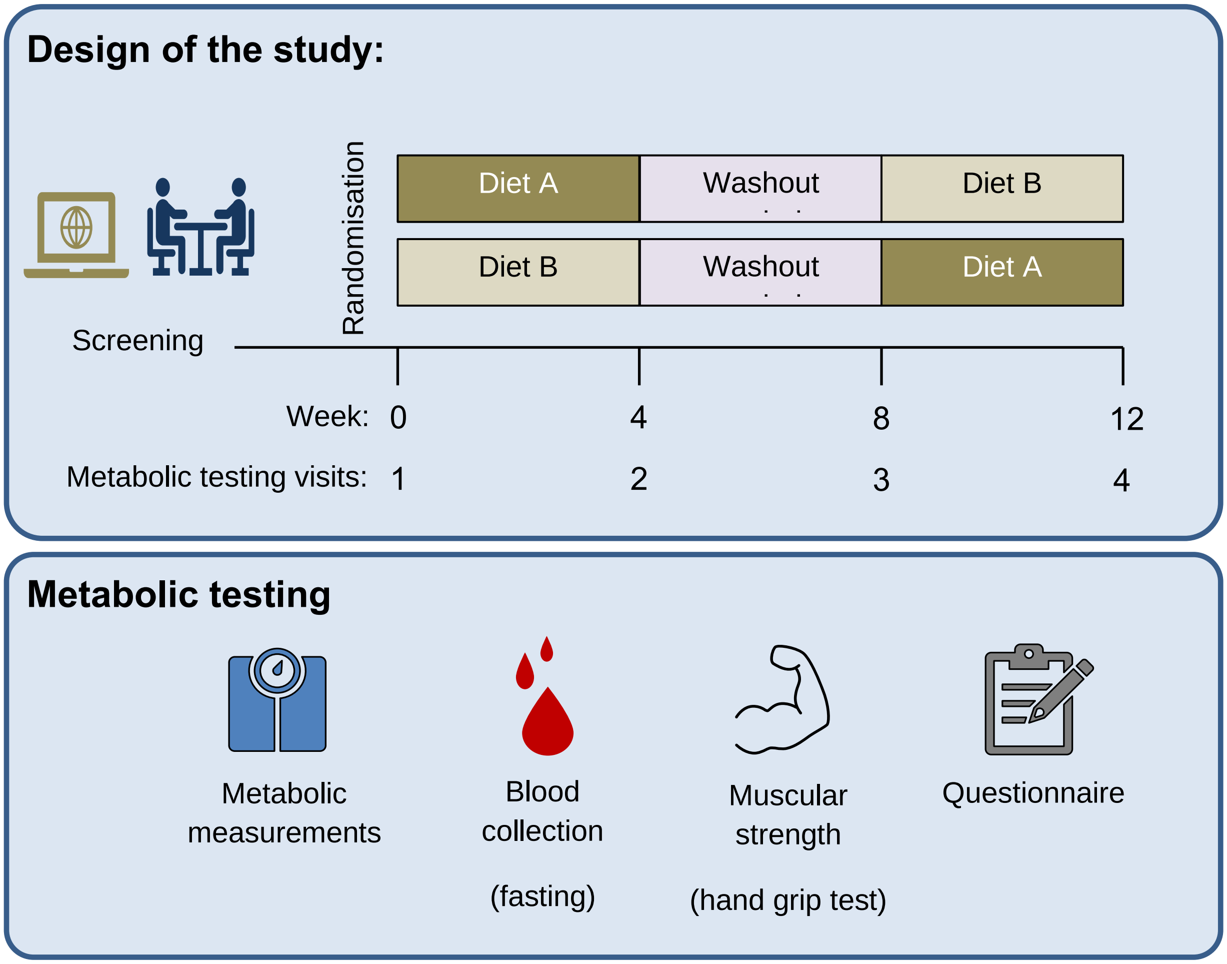


**Figure 1. Study Timeline**

**Diet Interventions (Study diets):**
During the two 4-week study diets, we will provide you with seven-day rotating menus and associated recipes, with the majority of foods in this menu provided to you either via supermarket home delivery or click & collect service. We will ask you to buy some highly perishable products like milk, fruits and vegetables at your own expense (you will have a choice of items to buy).

You will not know how the two study diets differ; this means you are blinded to which diet you are receiving at each timepoint. This is because we know that people can be influenced by diet composition in nutrition studies, and we are seeking to avoid any unconscious bias in this study.

In order to assist with blinding, we are keeping the food component of the diet as consistent as possible. We have incorporated the use of a supplemental powder that you will consume, most often as a drink, within your assigned diet. The supplemental powders will be used to change the macronutrient profile of the study diets rather than there being obvious changes in the food provided to you. The products used in the study powders are standard, nutritional supplements that are safe and widely available on the commercial market.

The two diets have been designed by the study dietitian, who is qualified to provide nutrition counselling. Both study diets will be healthy and balanced and chosen for you according to your height and body weight and whether you are male or female. We don't anticipate that you will lose or gain weight during the study. This study does not involve any fasting, apart from the overnight fast that is required prior to each metabolic visit. We will be able to take into account some dietary dislikes, which we will also ask you about at screening.

When you are following the study diets we will ask you to complete menu checklists daily, recording any food prescribed that was not eaten as well as any food eaten that was not prescribed. We ask that you be as honest as possible when completing these checklists. We will also send a general mid-diet check-in survey to give you the opportunity to express any concerns with following the diet. We also encourage you to contact the study team with concerns at any time as well.

**Washout period:**

During the 4-week washout period between the two diets, we will ask you to return to your usual habitual diet.

**How much time will my involvement in the project take?**

The time to participate in the study is 12 weeks in total. This includes the two 4-week study diets and the 4-week washout period described in Figure 1. You will spend approximately 7 hours in total at SAHMRI North Terrace, and the initial online screening questionnaires will take approximately 30 minutes to complete. In appreciation of the time commitment, travel and inconvenience, you will receive a $100 honorarium, in the form of a gift card, on completion of the study. You will also receive most of your foods free of charge during the two 4-week study diets, which is an estimated value of $800.

**Are there any risks associated with participating in this project?**

***Disclosing your information:***

Whilst all the information you provide to us will remain confidential, it may be subject to discovery in court or legal proceedings. This is a rare occurrence, but we are obligated to inform you of this risk. We will not disclose this information, or any information you provide to us, unless required by law.

*Blood samples*:
This test involves inserting a small needle into a vein in your arm. Inserting the needle can be associated with a small amount of pain and although complications are rare, it may occasionally cause bruising, localised bleeding, faintness and, in rare cases, infection. If you have ever experienced any of these complications in the past, bring them to the attention of the person performing the blood collection. If a blood sample is unable to be obtained by study staff at SAHMRI, you may be taken to SA Pathology at the Royal Adelaide Hospital to assist in obtaining a blood sample.

***Bioelectrical Impedance Analysis (BIA):***
This equipment measures muscle mass and body fat. You will change into a gown and stand barefoot on the base frame of the BIA equipment (like a set of scales with two handlebars). Your weight will be displayed on the screen and your body composition analysis measurement will take approximately 30 seconds. The BIA method uses a very low current which is negligible and will cause no harm to the human body. If you have any kind of contagious disease or injury on the palm of your hand/s or soles of your feet, then the BIA measurement cannot be performed.

**What are the potential benefits of the research project?**
This study is not directly assessing a treatment for a disease, and as such, you will not directly benefit from participating. This and future studies will inform us about whether using nutritional intervention targeting autophagy could potentially be useful to treat or prevent diseases that have been shown to relate to autophagy, such as cancer, Alzheimer's or cardiovascular disease. To date, researchers do not know whether autophagy-related measures may be indicators of your health; therefore, you will not be provided with your individual results, nor will we provide any counselling or advice regarding your results. Some of the other outcomes we measure (such as blood pressure) may be indicators of health. If this measure is outside of the normal range, you will be notified and provided with the relevant information to take to your GP for further evaluation.

**Can I withdraw from the project?**
Participation in this project is completely voluntary. If you agree to participate, you can withdraw from the study at any time. If you consent to future use of any remaining samples, you are free to withdraw your consent for future analysis by contacting the Principal Investigator at any time within the next 15 years. Your samples will be destroyed, and your data will be excluded from any analysis not already undertaken, and this will not affect your relationship with the University or SAHMRI now or in the future.

**What will happen to my information?**

*Confidentiality and privacy:*

At screening, you will be allocated a study ID, and all your information will be de-identified. All data collected will be stored on a password-protected database. While all efforts will be made to remove any information that might identify you, as the sample size is small, complete anonymity cannot be guaranteed. However, the utmost care will be taken to ensure that no personally identifying details are revealed.

*Storage:*

All data collected will be stored on a password protected database on a secure SAHMRI server. Any hard copy documents (where applicable) will be stored in a secure location within SAHMRI North Terrace. Your blood samples will be de-identified and stored for up to 15 years in freezers located on Level 6, SAHMRI, North Terrace.

*Publishing:*

Data collected from this study will be published (as summary data only) in scientific journals and presented at academic conferences. These disclosures will not identify you by name. At the end of the study, you will receive a summary report about the results of the study. This report will not identify you or any other participants by name.

*Sharing:*

Blood samples and data collected during the study may be shared with other researchers, in Australia or overseas, with your consent. These samples and/or data will be identified by your study ID only. After 15 years, samples will be disposed of in clinical waste bins and destroyed. Where your cultural sensitivities are relevant, specific agreements can be made with you and we will ensure that they are fulfilled when disposing of samples.

You will be asked whether you consent to be contacted for future research projects. We will only record your contact details on a password-protected file and contact you for future studies if you agree for us to do so.

Your information will only be used as described in this participant information sheet and it will only be disclosed according to the consent provided, except as required by law.

**Who do I contact if I have questions about the project?**
Should you have any questions or concerns before, during or after the study, please feel free to contact the SAHMRI Clinical Trials Platform on (08) 8128 4570 or by in the first instance.

The Principal Investigator, Dr Timothy Sargeant can be contacted on (08) 8128 4940 or by

**What if I have a complaint or any concerns?**
The study has been approved by the Human Research Ethics Committee at the University of Adelaide (Approval number H-2021-154). This research study will be conducted according to the NHMRC National Statement on Ethical Conduct in Human Research 2007 (Updated 2018). If you have questions or problems associated with the practical aspects of your participation in the study or wish to raise a concern or complaint about the study, please contact the Principal Investigator (Dr Timothy Sargeant, (08) 8128 4940).

If you wish to speak with an independent person regarding concerns or a complaint, the University's policy on research involving human participants, or your rights as a participant, please contact the Human Research Ethics Committee's Secretariat on:

Post: Level 4, Rundle Mall Plaza, 50 Rundle Mall, ADELAIDE SA 5000
Any complaint or concern will be treated in confidence and fully investigated. You will be informed of the outcome.

**If I want to participate, what do I do?**
In the first instance, please complete the online questionnaire available on the website or alternatively by contacting the research team by. The screening questionnaires will determine your eligibility for the study. If you would prefer to contact us by phone first, please call the ***SAHMRI Clinical Trials Platform on (08) 8128 4570*** and leave a message with your name and phone number if unable to speak to someone. A member of the study team will call you back to discuss the study with you and answer any questions you may have. If you are eligible to participate in the study, we will invite you to attend SAHMRI North Terrace for a screening visit.

Yours sincerely,

Dr Tim Sargeant and the NINA Study Team

### CONSENT FORM

**PROJECT TITLE:** Effect of **N**utritional **IN**terventions on **A**utophagy (The NINA study)

**HUMAN RESEARCH ETHICS COMMITTEE (HREC) APPROVAL NUMBER: H-2021-154**
**PRINCIPAL INVESTIGATOR:** Dr Timothy Sargeant

**1.** I have read the attached Information Sheet and agree to take part in the following research project:

| | |
|---|---|
| **Title:** | Effect of **N**utritional **IN**terventions on **A**utophagy (The **NINA** study) |
| **HREC Approval Number:** | **H-2021-154** |

**2.** I have had the project, so far as it affects me, and the potential risks and burdens fully explained to my satisfaction by the research worker. I have had the opportunity to ask any questions I may have about the project and my participation. My consent is given freely.

**3.** I have been given the opportunity to discuss the project with a friend or family member.

**4.** Although I understand the purpose of the research project is to improve the quality of health/medical care, it has also been explained that my involvement may not be of any benefit to me.

**5.** I agree to participate in the activities as outlined in the participant information sheet:
- Comply with the two (2) study diets and complete menu checklists daily for the duration of the study.
- Attend a screening visit followed by four (4) metabolic visits at SAHMRI during a 12-week period, which involve answering questions about my well-being and general health and undergoing health-related measurements (such as body weight and blood pressure).
- Provide a blood sample at each metabolic visit (x4).
- Undergo a Bioelectrical Impedance Analysis (BIA) to assess body composition at each metabolic visit.

**6.** I understand that my participation is voluntary and that I am free to withdraw my information from the project at any time.

**7.** Should I consent to future use of any blood samples, I understand that I am free to withdraw my consent for future analysis by contacting the principal researcher at any time within the next fifteen years. My samples will be destroyed, and my data will be excluded from any analysis not already undertaken.

**8.** I understand that if I decide not to take part or withdraw from the project, that this will not affect medical advice in the management of my health, now or in the future.

**9.** I have been informed that the information gained in the project may be published in a journal article, thesis, news article, conference presentations, website or report.

**10.** I have been informed that in the published materials I will not be identified, and my personal results will not be divulged.

**11.** I agree to my information being used for future research purposes as follows:

Research undertaken by these same researcher(s) Yes ☐ No ☐

Research undertaken by any researcher(s) Yes ☐ No ☐

**12.** I hereby provide 'extended' consent for the use of my data or tissue in future research projects that are:

- (i) an extension of, or closely related to, the original project: Yes ☐ No ☐
- (ii) in the same general area of research: Yes ☐ No ☐

**13.** I understand my information will only be disclosed according to the consent provided, except where disclosure is required by law.

**14.** I am aware that I should keep a copy of this Consent Form, when completed, and the attached Information Sheet.

**PLEASE COMPLETE THE SECTION BELOW:**

**Participant to complete:**

Name: _________________________ Signature: ____________________________ Date: _____________

**Researcher Declaration:**

I have described the nature of the research to this participant and in my opinion she/he understood the explanation.

Name of Researcher: ____________________Signature: _____________________

Position: ________________________ Date: _________________________________
